# Endocrine–metabolic network architecture reveals key bridge biomarkers in polycystic ovary syndrome

**DOI:** 10.64898/2026.07.10.26357756

**Authors:** Natalia Piórkowska, Grzegorz Franik, Anna Bizoń

**Affiliations:** Faculty of Information and Communication Technology, Wroclaw University of Science and Technology, Wrocław, Poland; Department of Endocrinological Gynecology, Faculty of Medical Sciences, Medical University of Silesia, Katowice, Poland; Department of Toxicology, Faculty of Pharmacy, Wroclaw Medical University, Wrocław, Poland

**Keywords:** Polycystic ovary syndrome, Network analysis, Graphical LASSO, Systems biology, Bridge biomarkers, Endocrine–metabolic interactions, Precision medicine, Biomarker networks

## Abstract

**Context:** Polycystic ovary syndrome (PCOS) is a heterogeneous endocrine disorder involving complex interactions among endocrine, metabolic, inflammatory, and thyroid pathways. However, the systems-level organization of these interactions remains poorly understood.

**Objective:** To reconstruct the endocrine–metabolic biomarker network in women with PCOS and identify bridge biomarkers integrating distinct physiological domains.

**Design:** Retrospective cross-sectional study.

**Setting:** Single tertiary referral center.

**Participants:** A total of 1,286 women diagnosed with PCOS according to the revised Rotterdam criteria.

**Methods:** Twenty-nine routinely measured laboratory biomarkers representing endocrine, metabolic, hematological/inflammatory, and thyroid domains were analyzed. Sparse Gaussian graphical models were estimated using Graphical LASSO with Extended Bayesian Information Criterion model selection. Network topology, node centrality, bridge centrality, bootstrap resampling, and predefined sensitivity analyses were performed.

**Results:** The reconstructed network comprised 29 biomarkers connected by 73 conditional dependency edges (network density, 0.18), demonstrating a modular but highly integrated endocrine–metabolic architecture. Conventional centrality analysis primarily identified biomarkers organizing local physiological modules, whereas bridge-centrality analysis revealed biomarkers coordinating communication between biological domains. Sex hormone–binding globulin exhibited the highest bridge strength, followed by fasting insulin, triglycerides, and high-density lipoprotein cholesterol. Additional reproducible bridge biomarkers included free thyroxine, white blood cell count, 2-hour plasma glucose, absolute neutrophil count, androstenedione, and anti-thyroglobulin antibodies. The leading bridge biomarkers remained stable across bootstrap resampling, complete-case reconstruction, and alternative network specifications.

**Conclusions:** PCOS is characterized by an integrated endocrine–metabolic network organized around a limited number of reproducible bridge biomarkers linking multiple physiological systems. Network analysis provides complementary systems-level information beyond conventional biomarker evaluation and may facilitate future biological phenotyping and precision medicine approaches in PCOS.

## 1. Introduction

Polycystic ovary syndrome (PCOS) is the most common endocrine disorder among women of reproductive age and is characterized by variable combinations of hyperandrogenism, ovulatory dysfunction, and polycystic ovarian morphology (PCOM) (1). Additionally is frequently accompanied by insulin resistance (IR), overweight/obesity, dyslipidemia, chronic low-grade inflammation, and an increased risk of metabolic and cardiovascular complications (2). The clinical presentation of PCOS is remarkably heterogeneous, reflecting substantial inter-individual variability in endocrine and metabolic disturbances (3).

Numerous circulating biomarkers reflecting reproductive, metabolic, inflammatory markers, hematological, and thyroid function have been investigated in PCOS, however these biomarkers have predominantly evaluated individually rather than as components of an integrated network (4). Although such reductionist approaches have substantially advanced our understanding of individual biomarkers and biological mechanisms, they may not adequately capture the complex network of interactions among multiple physiological systems that underlies the biological heterogeneity of PCOS. Increasing evidence indicates that endocrine, metabolic, immune, and thyroid pathways are highly interconnected and dynamically influence one another rather than acting independently (5,6).

PCOS is increasingly recognized as a complex systems disorder in which disease manifestations emerge from interactions among multiple biological networks rather than isolated abnormalities (7). In this context, systems biology approaches have gained considerable interest because they enable the investigation of coordinated relationships among biomarkers and may identify key regulatory components underlying disease heterogeneity. Network analysis represents one such approach, allowing biomarkers to be analyzed as interconnected nodes linked through conditional dependencies after accounting for the influence of all remaining variables. Unlike conventional correlation analyses, network models identify direct statistical associations and provide insight into the overall organization of biological systems.

Among contemporary network approaches, Graphical Gaussian Models (GGMs), estimated using Graphical LASSO regularization, are widely used to infer sparse biological networks from high-dimensional omics datasets by identifying conditional dependencies among variables (8,9). These models reduce spurious associations while preserving the strongest conditional relationships between variables. Furthermore, graph-theoretical measures such as node and bridge centrality facilitate the identification of biomarkers that either occupy influential positions within functional modules or serve as bridges linking distinct physiological domains (10,11). In particular, bridge biomarkers may represent key integrative regulators linking endocrine, metabolic, inflammatory, hematological, and thyroid systems, thereby providing biologically meaningful insight beyond conventional biomarker comparisons.

Despite growing interest in system biology, network-based investigations of PCOS remain limited, particularly those integrating endocrine, metabolic, inflammatory, hematological, and thyroid biomarkers within a single analytical framework. Consequently, the organizational structure of these interconnected biological systems and the biomarkers coordinating communication between them remain poorly understood.

Therefore, the aim of the present study was to reconstruct the integrated endocrine-metabolic network in young women with PCOS using Graphical LASSO network analysis and to identify central and bridge biomarkers that coordinate interactions endocrine, metabolic, inflammatory, hematological, and thyroid physiological domains. In addition, we evaluated the robustness of the identified network architecture using multiple complementary sensitive analyses to assess the reproducibility of the observed biological relationships.

## 2. Methods

### 2.1. Study design and objectives

This retrospective cross-sectional study was designed to characterize the biological architecture of PCOS using a systems-level network approach integrating endocrine, metabolic, inflammatory, and thyroid biomarkers.

Rather than evaluating isolated associations between individual biomarkers, the study aimed to reconstruct the conditional dependency network underlying PCOS biology and to identify biomarkers occupying central and bridge positions within this multidimensional system. The analytical framework was based on sparse Gaussian graphical models, which estimate direct conditional relationships between biomarkers while accounting for the influence of all remaining variables in the network.

The primary objective was to identify bridge biomarkers connecting endocrine, metabolic, inflammatory, and thyroid biological domains. Bridge biomarkers were defined as variables exhibiting the highest connectivity between predefined physiological domains and were considered potential integrators of multiple biological systems involved in PCOS.

The secondary objectives were to: (i) characterize the global topology of the endocrine–metabolic biomarker network; (ii) quantify node importance using complementary graph-theoretical centrality measures; (iii) evaluate the robustness and reproducibility of the inferred network through bootstrap resampling; and (iv) assess the stability of bridge biomarkers across alternative network regularization parameters and model-selection strategies.

The study was hypothesis-driven. We hypothesized that PCOS exhibits a modular biological architecture composed of interconnected endocrine, metabolic, inflammatory, and thyroid subnetworks rather than a single homogeneous biological phenotype. Furthermore, we hypothesized that a limited number of biomarkers function as bridge nodes linking these physiological domains, thereby organizing the overall biological network architecture.

No supervised prediction models were developed. All analyses focused exclusively on network estimation, topological characterization, and robustness assessment of the inferred biological architecture.

### 2.2. Study population

This retrospective cross-sectional study included women diagnosed with PCOS who underwent comprehensive endocrine and metabolic evaluation at the Department of Endocrinological Gynecology, Medical University of Silesia, Katowice, Poland, between January 2018 and May 2025.

PCOS was diagnosed according to the revised Rotterdam criteria (12), requiring the presence of at least two of the following features after exclusion of alternative etiologies: (i) oligo- or anovulation, (ii) clinical and/or biochemical hyperandrogenism, and (iii) polycystic ovarian morphology on ultrasonography.

As part of routine clinical assessment, all participants underwent standardized clinical examination, fasting laboratory testing, and metabolic evaluation according to the institutional diagnostic protocol. Laboratory investigations included reproductive hormones, thyroid function tests, glucose metabolism, lipid profile, complete blood count parameters, and additional endocrine biomarkers routinely collected during diagnostic work-up.

Women with incomplete diagnostic evaluation preventing confirmation of the PCOS diagnosis were excluded. Additional exclusion criteria comprised endocrine disorders capable of explaining hyperandrogenism or ovulatory dysfunction, including non-classic congenital adrenal hyperplasia, Cushing syndrome, androgen-secreting tumors, hyperprolactinemia, and overt thyroid dysfunction diagnosed before establishment of the PCOS diagnosis.

The source cohort consisted of 1,286 women with confirmed PCOS, representing all eligible participants who fulfilled the study eligibility criteria during the study period. Because laboratory investigations were performed as part of routine clinical practice, the availability of individual biomarkers varied across participants, resulting in variable patterns of missing data across laboratory variables.

To maximize statistical power while preserving the complete study cohort, the primary network analyses were performed using a harmonized analytical dataset generated after predefined median imputation of missing laboratory measurements. Median imputation was selected because it provides a robust, distribution-preserving approach for routinely collected clinical biomarkers and minimizes the influence of extreme observations. The complete-case dataset was retained exclusively for predefined sensitivity analyses evaluating the influence of missing-data handling on network topology, centrality measures, and bridge biomarker identification.

The derivation of the analytical datasets used in the primary and sensitivity analyses is summarized in the study flow diagram.

The study was conducted in accordance with the Declaration of Helsinki and was approved by the Bioethical Committee of Wroclaw Medical University (approval No. 254/2021). All clinical data were analyzed in anonymized form.

### 2.3. Clinical and laboratory assessment

All clinical and laboratory measurements were obtained as part of routine diagnostic evaluation performed according to the institutional PCOS assessment protocol. Clinical data were extracted retrospectively from the institutional electronic medical records, whereas laboratory measurements were retrieved from the certified laboratory information system.

Blood samples were collected in the morning after an overnight fast. Whenever clinically feasible, hormonal measurements were performed during the early follicular phase of the menstrual cycle (cycle days 2–5) or after spontaneous or progesterone-induced withdrawal bleeding in women with oligo- or amenorrhea, following routine clinical practice.

Laboratory analyses were performed in the Central Laboratory of the University Clinical Centre of the Medical University of Silesia (Katowice, Poland), an accredited clinical laboratory operating under standardized internal and external quality-control procedures. Hormonal measurements, including luteinizing hormone (LH), follicle-stimulating hormone (FSH), total testosterone, free testosterone, sex hormone-binding globulin (SHBG), dehydroepiandrosterone sulfate (DHEAS), androstenedione (ANDRO), anti-Müllerian hormone (AMH), thyroid-stimulating hormone (TSH), free thyroxine (fT4), anti-thyroid peroxidase antibodies (anti-TPO), anti-thyroglobulin antibodies (anti-Tg), cortisol, and other endocrine biomarkers, were determined using standardized electrochemiluminescence immunoassays (ECLIA; Roche Diagnostics, Mannheim, Germany). Routine biochemical measurements, including fasting and 2-hour plasma glucose during a 75-g oral glucose tolerance test (OGTT), fasting insulin, total cholesterol (TCHOL), high-density lipoprotein cholesterol (HDL-C), low-density lipoprotein cholesterol (LDL-C), triglycerides (TG), C-reactive protein (CRP), and additional biochemical parameters, were measured using standardized automated enzymatic methods implemented in routine clinical diagnostics.

Complete blood count (CBC) parameters, including leukocyte differential counts, erythrocyte indices, platelet indices, and absolute leukocyte subpopulations, were measured using automated hematology analyzers according to the manufacturer’s recommendations and laboratory quality-control procedures.

All laboratory measurements were performed using routine clinical assays throughout the study period. Laboratory methods remained consistent with the diagnostic standards implemented at the participating institution, ensuring comparability of measurements across the study cohort.

Only variables representing routinely collected clinical biomarkers were considered for subsequent analyses. No additional experimental assays were performed specifically for the present study.

### 2.4. Network variable selection

The primary biomarker network was constructed using a predefined set of routinely measured laboratory biomarkers selected *a priori* on the basis of their established biological relevance to the pathophysiology of PCOS, routine availability within the institutional diagnostic protocol, and suitability for multivariable network modeling. Variable selection was completed before any network estimation or graph-theoretical analyses were performed.

To minimize circularity and avoid data-driven feature selection, biomarkers were selected according to their physiological role rather than statistical criteria such as univariate associations, pairwise correlations, or network topology. Only primary laboratory measurements representing distinct biological processes were included in the primary network.

The primary network consisted of 29 biologically independent laboratory biomarkers, grouped into four predefined physiological domains.

(i) The endocrine domain comprised biomarkers reflecting hypothalamic–pituitary–ovarian and adrenal function: LH, FSH, SHBG, DHEAS, androstenedione (ANDRO), AMH, morning cortisol, and evening cortisol.
(ii) The metabolic domain included fasting plasma glucose, 2-hour plasma glucose during the oral glucose tolerance test (OGTT), fasting insulin, total cholesterol, LDL-C, HDL-C, and TG.
(iii) The hematological and inflammatory domain included total white blood cell count (WBC), absolute neutrophil count, absolute lymphocyte count, absolute monocyte count, platelet count, red cell distribution width (RDW), mean platelet volume (MPV), platelet distribution width (PDW), and platelet large cell ratio (P-LCR).
(iv) The thyroid domain comprised TSH, fT4, anti-TPO, and anti-Tg.

Variables representing deterministic mathematical transformations of primary laboratory measurements—including the LH/FSH ratio, free androgen index (FAI), homeostatic model assessment for insulin resistance (HOMA-IR), quantitative insulin sensitivity check index (QUICKI), triglyceride–glucose (TyG) index, triglyceride-to-HDL cholesterol ratio (TG/HDL-C), non-HDL cholesterol, neutrophil-to-lymphocyte ratio (NLR), platelet-to-lymphocyte ratio (PLR), systemic immune-inflammation index (SII), lymphocyte-to-monocyte ratio (LMR), and cortisol ratio—were intentionally excluded from the primary network because they represent deterministic combinations of primary variables. Including both primary and derived biomarkers within the same network may introduce mathematical coupling, artificially increase conditional dependencies, and bias estimation of the underlying biological network architecture.

These composite indices were evaluated separately in predefined sensitivity analyses to determine whether their inclusion altered global network topology, node-centrality measures, or identification of bridge biomarkers.

Consequently, the primary network was intentionally restricted to biologically independent laboratory measurements, enabling estimation of conditional dependencies that more closely reflected physiological interactions rather than mathematical relationships among derived variables.

### 2.5. Data harmonization and preprocessing

All preprocessing procedures were predefined before network estimation and implemented within a fully reproducible computational pipeline. The objective of preprocessing was to generate a harmonized analytical dataset while minimizing investigator-dependent decisions and preserving biological interpretability.

Clinical and laboratory variables were extracted from the institutional database and harmonized using predefined variable-mapping rules. Measurements representing the same biological parameter but recorded under different laboratory names, abbreviations, or reporting formats were consolidated into standardized feature identifiers before further analyses.

Quality-control procedures included verification of variable identity, assessment of biologically plausible measurement ranges, evaluation of missing-data patterns, and identification of inconsistencies in laboratory coding. Variables with ambiguous biological interpretation or inconsistent measurement definitions were excluded before network construction. No variables were selected or removed on the basis of network topology, correlation structure, or graph-theoretical results.

Laboratory values reported as character strings, interval ranges, or threshold expressions (e.g., “<x” or “>x”) were converted into numeric representations using predefined parsing rules. All unit harmonization procedures were completed before statistical analyses to ensure comparability across the study cohort.

Continuous variables were inspected for distributional characteristics and standardized using z-score normalization prior to network estimation. Standardization was performed to place biomarkers measured on different numerical scales onto a common metric without altering their underlying correlation structure.

No univariate filtering based on statistical significance, variance, or pairwise correlation was performed before network estimation. This approach ensured that network topology emerged from multivariable conditional dependency modeling rather than from investigator-driven feature reduction.

Derived clinical indices were generated automatically within the preprocessing pipeline when required for predefined sensitivity analyses but were intentionally excluded from construction of the primary biomarker network, as described in the previous section.

The complete preprocessing workflow was deterministic and fully reproducible. Intermediate datasets generated at each computational stage were automatically stored and served as the exclusive input for subsequent analytical steps, ensuring complete traceability of the entire network analysis pipeline.

### 2.6. Missing data handling

Missing laboratory measurements were expected because the study was based on routinely collected clinical data, and not all biomarkers were requested for every participant during standard diagnostic evaluation. Consequently, the pattern of missingness differed across laboratory variables according to clinical decision-making rather than the objectives of the present study.

To preserve the full source cohort and maximize statistical power for multivariable network estimation, the primary analyses were performed using a harmonized analytical dataset generated after missing-value imputation. Missing values were imputed independently for each biomarker using median imputation implemented through the SimpleImputer algorithm from the *scikit-learn* library. Median imputation was selected because it is robust to skewed laboratory distributions, preserves observed measurement ranges, and minimizes the influence of extreme values commonly encountered in routine clinical biomarkers.

Prior to imputation, the extent and distribution of missing values were examined descriptively for all candidate biomarkers. Patterns of missingness were evaluated as part of the quality-control workflow to identify variables with limited availability or systematic missing-data patterns that could influence downstream analyses. These assessments were performed solely for quality assurance and did not influence biomarker selection, which had been predefined *a priori*.

The median-imputed dataset served as the primary analytical dataset for network estimation, Graphical LASSO model construction, centrality analyses, and bridge-centrality analyses. This approach enabled inclusion of all 1,286 eligible women while maintaining a consistent covariance structure across all biomarkers included in the network.

To evaluate the influence of missing-data handling on the inferred biological architecture, a predefined complete-case analysis was performed as a sensitivity analysis. Networks reconstructed from the complete-case dataset were compared with the primary imputed network with respect to global network topology, edge preservation, node-centrality measures, bridge-centrality rankings, and identification of the highest-ranking bridge biomarkers.

Agreement between the primary and sensitivity analyses was interpreted as evidence that the principal biological findings were robust to alternative strategies for handling missing data. The complete-case analysis therefore served exclusively as an assessment of robustness and was not used for primary biological inference.

### 2.7. Construction of the primary biomarker network

The primary biomarker network was constructed to represent the conditional dependency structure among routinely measured laboratory biomarkers in women with polycystic ovary syndrome. Each biomarker was represented as a node, whereas edges represented direct statistical associations estimated after conditioning on all remaining variables included in the network.

Only the predefined primary biomarkers described in the previous section were included in construction of the primary network. Variables representing deterministic mathematical transformations of other measurements were intentionally excluded from the primary network because their inclusion together with their constituent variables may introduce mathematical coupling and distort estimation of conditional dependency structure.

Prior to network estimation, continuous variables were standardized using z-score normalization. Standardization was performed solely to place biomarkers measured on different numerical scales onto a common metric and did not alter the underlying correlation structure among variables.

Network construction was based on Gaussian graphical modeling, in which conditional dependencies between biomarkers are represented by the non-zero elements of the precision, or inverse covariance, matrix. Unlike conventional correlation networks, Gaussian graphical models estimate direct relationships between biomarkers while accounting for all remaining variables in the network, thereby reducing indirect associations arising from shared covariance.

The primary network was estimated using the median-imputed analytical dataset comprising all 1,286 eligible women with PCOS. The resulting graph was undirected, with nodes corresponding to individual biomarkers and edges representing non-zero conditional dependencies identified after regularized precision-matrix estimation.

Biological domains—endocrine, metabolic, hematological and inflammatory, and thyroid—were predefined before network construction and used exclusively for biological interpretation and subsequent bridge-centrality analyses. Domain membership did not influence estimation of network edges or network topology.

The primary network represented the prespecified analytical model of the study. Complete-case network reconstruction and complementary networks incorporating derived clinical indices were generated only as predefined sensitivity analyses to evaluate whether missing-data handling or inclusion of composite biomarkers altered network topology or the identification of central and bridge biomarkers.

### 2.8. Graphical LASSO estimation and model selection

The primary biomarker network was estimated using the graphical least absolute shrinkage and selection operator (Graphical LASSO), a regularized Gaussian graphical modeling approach that estimates a sparse inverse covariance (precision) matrix. This framework identifies direct conditional dependencies between biomarkers while accounting for all remaining variables included in the model, thereby reducing indirect associations arising from shared covariance.

Network sparsity was achieved through L1 regularization applied to the precision matrix. Increasing the regularization parameter progressively removes weak conditional dependencies, resulting in a more parsimonious and biologically interpretable network while reducing the influence of spurious associations.

Selection of the optimal regularization parameter followed a predefined model-selection strategy established before biological interpretation. Candidate models were estimated across a grid of regularization parameters and evaluated using the Extended Bayesian Information Criterion (EBIC), which balances model fit against network complexity in high-dimensional settings. To avoid biologically implausible overconnected networks, EBIC optimization was supplemented by a predefined network density constraint of ≤0.20, ensuring that the final network remained sufficiently sparse for meaningful biological interpretation.

The final Graphical LASSO model satisfied both predefined selection criteria and was obtained using a regularization parameter of α = 0.125. The resulting network contained 29 biomarkers connected by 73 conditional dependency edges, corresponding to a network density of 0.18.

The selected precision matrix was transformed into an undirected weighted network in which non-zero off-diagonal elements represented conditional dependencies between biomarker pairs after adjustment for all remaining biomarkers included in the model. Edge weights reflected the magnitude of partial associations, whereas zero-valued elements indicated conditional independence.

To evaluate the robustness of network estimation, complementary Graphical LASSO models were reconstructed across neighboring regularization parameters and alternative sparsity levels. Global network topology, edge preservation, node-centrality measures, and bridge-centrality rankings were compared across candidate models to assess whether the principal biological findings remained stable despite moderate variation in model regularization.

Model selection was completed before computation of centrality measures or biological interpretation. Consequently, no modifications to the selected network were made after inspection of node rankings, bridge-centrality measures, or downstream biological results.

### 2.9. Network topology and centrality analysis

Following network estimation, the topological properties of the inferred biomarker network were characterized using graph-theoretical measures describing both global network organization and node-specific importance.

Global network topology was evaluated by quantifying the number of nodes, number of edges, network density, average node degree, connected components, clustering coefficient, and average shortest path length where applicable. These measures were used to describe the overall structural organization of the endocrine–metabolic biomarker network rather than for model selection.

The relative importance of individual biomarkers within the network was assessed using complementary node-centrality measures. Node strength was calculated as the sum of the absolute weights of all edges connected to a given biomarker and was considered the primary measure of overall network influence. In addition, betweenness centrality was calculated to quantify the extent to which a biomarker participated in the shortest communication paths linking different regions of the network, whereas closeness centrality reflected the average topological distance between a biomarker and all remaining nodes.

Because individual centrality measures capture different aspects of network organization, biomarkers were not ranked according to a single metric alone. Instead, centrality profiles were interpreted jointly to distinguish biomarkers exhibiting consistently high topological importance across multiple graph-theoretical measures from those demonstrating isolated prominence within a single metric.

Centrality measures were calculated from the weighted undirected network estimated by the Graphical LASSO model. Edge weights were retained during computation of node strength, whereas shortest-path-based metrics incorporated weighted network distances derived from the estimated partial associations.

To facilitate biological interpretation, node-centrality values were visualized using ranked centrality plots and network diagrams in which node size was proportional to overall centrality. These visualizations were generated after completion of all network estimation procedures and served exclusively to summarize the inferred biological architecture.

The identification of bridge biomarkers was not based on conventional node-centrality measures alone. Bridge centrality was evaluated separately using predefined biological domains, as described in the following section.

### 2.10. Bridge centrality analysis

The primary endpoint of the study was the identification of bridge biomarkers connecting the predefined biological domains represented within the endocrine–metabolic network. Bridge-centrality analysis was performed to identify biomarkers preferentially linking distinct physiological systems rather than biomarkers exhibiting high connectivity only within a single biological domain.

Before network estimation, each biomarker was assigned a priori to one of four predefined biological domains: endocrine, metabolic, hematological and inflammatory, or thyroid. Domain assignment was based exclusively on established physiological function and routine clinical interpretation and was completed before network reconstruction. Domain membership remained fixed throughout all analyses and was not modified according to the observed network topology.

Bridge importance was quantified using bridge strength centrality, defined as the sum of the absolute weights of all edges connecting a biomarker to nodes belonging to the other predefined biological domains. Consequently, bridge-strength values reflected the extent to which a biomarker integrated distinct physiological systems rather than its connectivity within its own biological domain.

Bridge-centrality measures were calculated with respect to the predefined four-domain network structure and were evaluated independently of conventional node-centrality metrics. Whereas node strength quantifies overall network connectivity, bridge strength specifically measures inter-domain connectivity and therefore identifies biomarkers that potentially coordinate communication between biologically distinct functional modules.

Bridge biomarkers were identified using the continuous distribution of bridge-strength values rather than predefined threshold-based classification. Accordingly, all biomarkers contributed to the bridge-centrality ranking, whereas biological interpretation focused on biomarkers consistently occupying the highest positions in the bridge-strength ranking. No arbitrary cut-off values were applied to define bridge biomarkers.

To facilitate biological interpretation, bridge-strength rankings were interpreted together with conventional node-centrality measures. This complementary approach enabled distinction between biomarkers exhibiting high global network influence and those functioning primarily as integrators of multiple physiological domains.

The robustness of bridge-centrality estimates was evaluated using bootstrap resampling, alternative Graphical LASSO regularization parameters, and complementary network specifications. Biomarkers consistently demonstrating high bridge-strength rankings across these predefined sensitivity analyses were considered robust bridge biomarkers.

Bridge-centrality analysis constituted the primary biological interpretation of the inferred network architecture and was performed only after completion of network estimation and model selection. Results of the bridge-centrality analysis did not influence any preceding stage of variable selection, network estimation, or model optimization.

### 2.11. Network robustness and sensitivity analyses

A series of predefined robustness and sensitivity analyses was performed to evaluate the stability of the inferred biological network and to determine whether the identification of central and bridge biomarkers depended on specific analytical assumptions.

First, the influence of missing-data handling was assessed by comparing the primary median-imputed network with networks reconstructed from the complete-case dataset. Global network topology, edge preservation, node-centrality measures, and bridge-centrality rankings were compared between both analytical strategies to evaluate the consistency of the inferred biological architecture.

Second, the stability of network estimation was evaluated across alternative regularization parameters. Networks were reconstructed over a predefined range of regularization strengths surrounding the EBIC-selected solution. Global network characteristics, including the number of edges, network density, connectedness, and node-centrality rankings, were compared to determine whether the principal biological findings remained consistent under different levels of network sparsity.

Third, additional sensitivity analyses examined the influence of network-density constraints during model selection. Candidate networks satisfying alternative predefined density thresholds were evaluated to assess whether the principal bridge biomarkers remained stable despite moderate variations in overall network complexity.

Fourth, the effect of variable composition was assessed by reconstructing complementary networks that included clinically established derived biomarkers, including composite endocrine, metabolic, and hematological/inflammatory indices. These analyses were performed exclusively as sensitivity analyses to determine whether inclusion of deterministic composite variables altered the overall biological interpretation obtained from the primary network consisting exclusively of primary laboratory measurements.

The robustness of node-centrality and bridge-centrality estimates was further evaluated using nonparametric bootstrap resampling with 500 iterations. For each bootstrap sample, the complete network estimation procedure—including regularized precision-matrix estimation, network reconstruction, and computation of graph-theoretical measures—was repeated independently. Bootstrap distributions were subsequently used to evaluate the reproducibility of node rankings and the stability of bridge biomarker identification.

Biomarkers consistently demonstrating high bridge-centrality rankings across bootstrap resampling, alternative regularization parameters, and complementary network specifications were considered robust bridge biomarkers. Conversely, biomarkers exhibiting marked variability across sensitivity analyses were interpreted cautiously and regarded as exploratory findings.

Sensitivity analyses were specified before biological interpretation and were not used to modify the primary network or redefine the principal study conclusions. Instead, they served to evaluate the reproducibility, robustness, and generalizability of the inferred endocrine–metabolic network architecture.

### 2.12. Statistical analysis

Statistical analyses were conducted according to a predefined analytical protocol established before network reconstruction and biological interpretation. Descriptive statistics were used to summarize the clinical and laboratory characteristics of the study cohort and to evaluate data quality prior to network estimation.

Continuous variables were summarized as mean ± standard deviation (SD) or median with interquartile range (IQR), depending on their distribution. Categorical variables were presented as counts and percentages.

The primary analytical dataset consisted of the median-imputed cohort including all **1,286** eligible women with PCOS. This dataset was used for all primary analyses, including Graphical LASSO network estimation, network topology assessment, node-centrality analysis, bridge-centrality analysis, and biological interpretation. Complete-case analyses were performed exclusively as predefined sensitivity analyses to evaluate the influence of missing-data handling on the inferred network architecture.

The primary endpoint of the study was the identification of bridge biomarkers connecting the predefined endocrine, metabolic, hematological and inflammatory, and thyroid biological domains. Secondary endpoints included characterization of global network topology, identification of highly connected biomarkers using complementary node-centrality measures, and assessment of network robustness across predefined sensitivity analyses.

Graph-theoretical measures were interpreted as continuous descriptors of network organization rather than hypothesis-testing statistics. Accordingly, node-centrality and bridge-centrality measures were evaluated using their relative rankings, and biological interpretation focused on biomarkers demonstrating consistently high rankings across complementary analytical approaches. No arbitrary threshold values were used to classify biomarkers as central or bridge nodes.

The robustness of the inferred network architecture was evaluated by comparing results obtained from the primary median-imputed dataset with predefined sensitivity analyses, including complete-case network reconstruction, alternative Graphical LASSO regularization parameters, predefined network-density constraints, complementary network specifications, and nonparametric bootstrap resampling.

Where conventional statistical hypothesis testing was performed outside the network estimation framework, all tests were two-sided, and a *P* value <0.05 was considered statistically significant. Adjustment for multiple comparisons was performed using the Benjamini–Hochberg false discovery rate (FDR) procedure whenever applicable. Effect estimates were accompanied by 95% confidence intervals where appropriate.

### 2.13. Software and reproducibility

All computational analyses were performed in Python using a fully reproducible analytical pipeline composed of sequential computational modules developed before biological interpretation. The complete workflow was implemented as a series of independent Google Colaboratory notebooks, each corresponding to a predefined stage of the analytical pipeline, including data harmonization, preprocessing, missing-data handling, network construction, Graphical LASSO estimation, network topology analysis, bridge-centrality analysis, robustness assessment, sensitivity analyses, and final quality-control procedures.

The analytical workflow was strictly sequential and deterministic. Outputs generated by each notebook—including harmonized datasets, quality-control reports, intermediate analytical tables, estimated network objects, graphical visualizations, model-selection summaries, and statistical reports—were automatically saved to dedicated output directories and served as the exclusive input for the subsequent analytical stage. This modular architecture ensured complete traceability of every analytical decision from the raw clinical dataset to the final reported results.

All analyses were performed using open-source scientific computing libraries, including NumPy, pandas, SciPy, scikit-learn, NetworkX, Matplotlib, and related Python packages. Sparse Gaussian graphical models were estimated using Graphical LASSO implementations available within the Python scientific ecosystem, whereas graph-theoretical measures and bridge-centrality analyses were computed using established network-analysis libraries. Exact software versions, package dependencies, computational environment specifications, and pipeline configuration files were automatically recorded during execution to ensure computational reproducibility.

Randomized procedures, including bootstrap resampling, were performed using predefined random seeds. All preprocessing decisions, variable-selection rules, model-selection criteria, regularization procedures, and sensitivity analyses were specified before biological interpretation and executed automatically without manual modification of intermediate analytical results.

The computational pipeline was designed according to the principles of reproducible research. All executable notebooks, source code, configuration files, intermediate outputs, documentation, and metadata describing the analytical workflow will be made publicly available through a version-controlled GitHub repository upon publication. The repository will enable independent reproduction of all analyses, verification of reported findings, and reuse of the complete analytical framework for future studies investigating biological network architecture in endocrine and metabolic disorders.

## Results

### 3.1 Study cohort

The source cohort comprised 1,286 women with confirmed PCOS who fulfilled all study eligibility criteria and underwent routine endocrine and metabolic evaluation during the study period.

Because laboratory investigations were performed according to routine clinical practice, individual biomarkers exhibited variable proportions of missing observations. To preserve the entire study cohort and maximize statistical power for multivariable network estimation, the primary analyses were performed using the predefined median-imputed analytical dataset including all 1,286 participants. A complete-case dataset was reconstructed separately and used exclusively for predefined sensitivity analyses evaluating the influence of missing-data handling on the inferred network architecture.

The primary network was constructed from 29 predefined laboratory biomarkers representing four biological domains: endocrine, metabolic, hematological and inflammatory, and thyroid. These biomarkers were selected *a priori* before network estimation and constituted the prespecified analytical framework for all subsequent analyses.

Figure 1 Study workflow illustrating participant inclusion, data harmonization, preprocessing, primary biomarker selection, generation of the median-imputed analytical dataset used for the primary analyses, and the complete-case dataset used for predefined sensitivity analyses.

**Figure 1.**
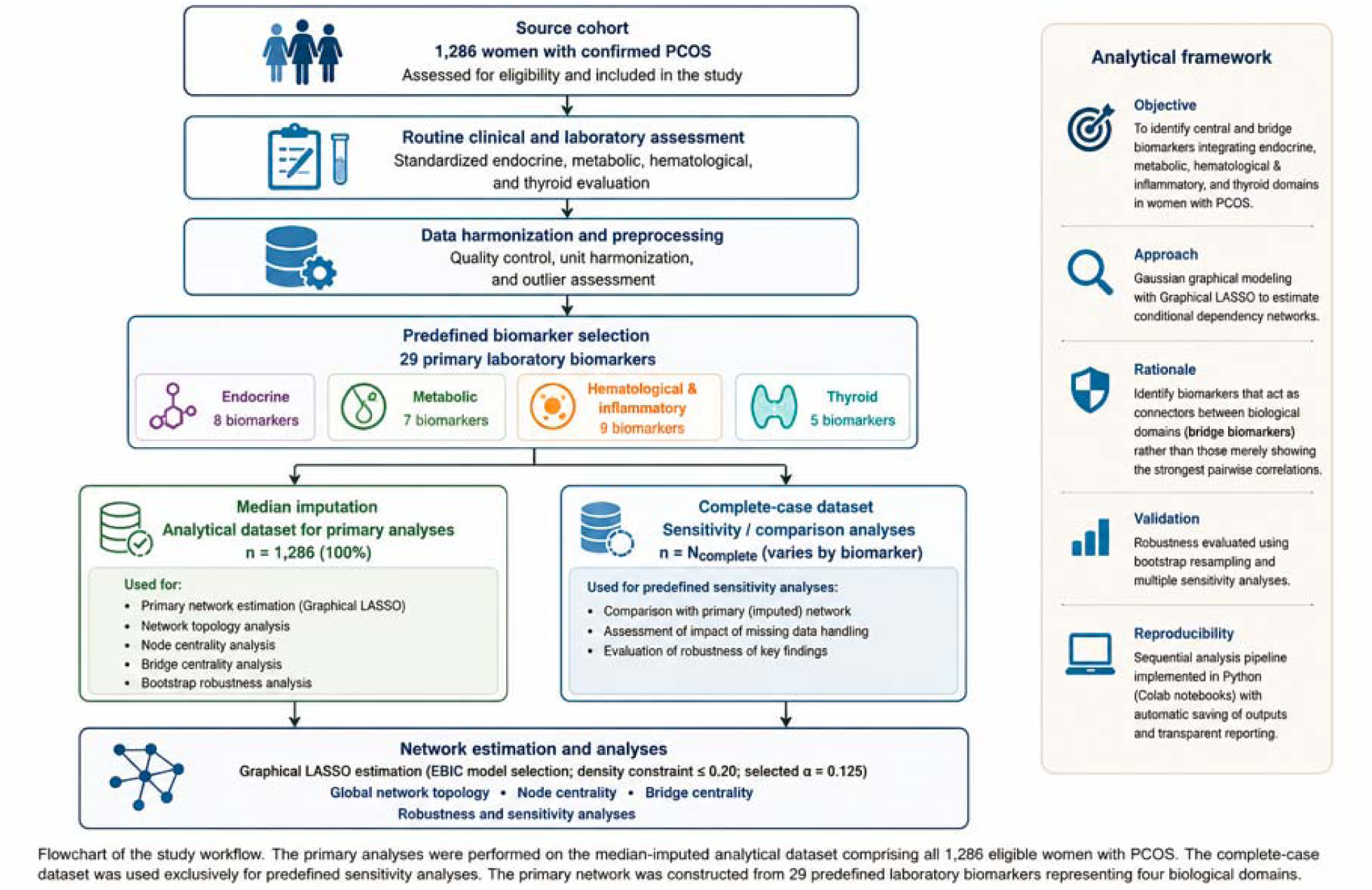
Study workflow and derivation of the analytical datasets used for network analysis

### 3.2 Global network architecture

The reconstructed endocrine–metabolic network revealed a sparse but highly interconnected biological architecture linking endocrine, metabolic, hematological and inflammatory, and thyroid biomarkers into a single integrated system. Rather than forming isolated clusters, biomarkers from different physiological domains were connected through multiple conditional dependencies, indicating extensive cross-talk between endocrine regulation, metabolic homeostasis, immune function, and thyroid physiology in women with PCOS.

Application of the predefined Graphical LASSO model-selection strategy resulted in a network comprising 29 biomarkers connected by 73 conditional dependency edges, corresponding to a network density of 0.18. The selected regularization parameter (α = 0.125) satisfied both predefined optimization criteria, including the minimum Extended Bayesian Information Criterion (EBIC) and the prespecified network-density constraint (≤0.20), yielding a parsimonious and biologically interpretable network.

Figure 2 illustrates the inferred global network architecture. Distinct biological domains remained visually recognizable; however, numerous inter-domain connections demonstrated that endocrine, metabolic, hematological and inflammatory, and thyroid processes did not operate as independent physiological modules. Instead, the network exhibited an integrated organization characterized by extensive conditional interactions across biological systems.

**Figure 2.**
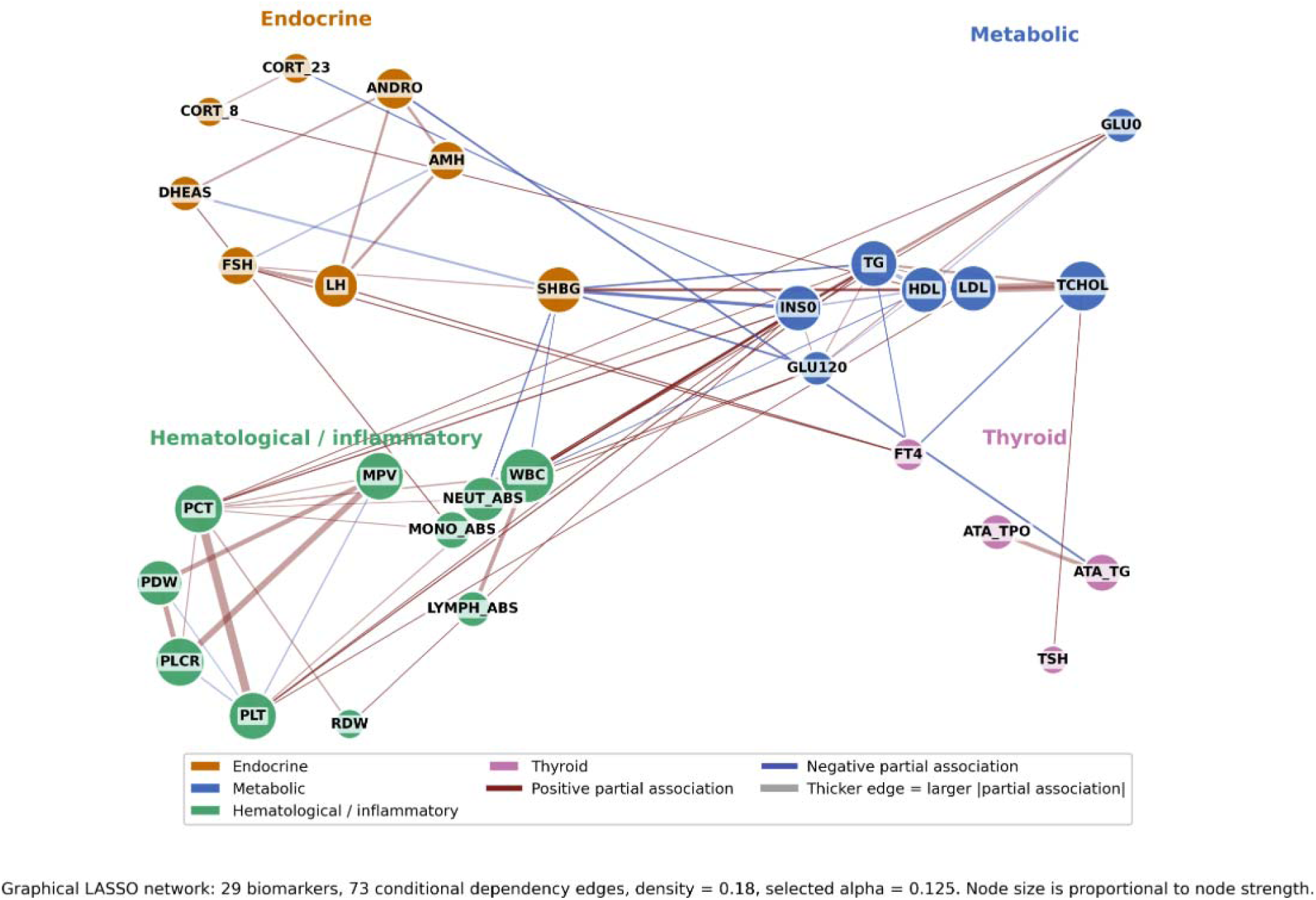
Global endocrine–metabolic biomarker network in women with PCOS Legend: LH-luteinizing hormone; FSH- follicle-stimulating hormone; DHEA-S-dehydroepiandrosterone sulfate; AMH - anti-Müllerian hormone; ANDRO - androstendione; CORT_8 - cortisol at 8 a.m.; CORT_23 - cortisol at 1 p.m.; SHBG - sex hormone-binding globulin; HDL-C - high density lipoprotein cholesterol; LDL- C - low density lipoprotein cholesterol; TCHOL - total cholesterol; TG - triglycerides; GLU0 - fasting glucose; GLU120 - glucose after OGTT; INS0 - fasting insulin; TSH - thyroid-stimulating hormone; fT4 - free thyroxine; ATA_TPO - thyroid peroxidase antibodies; ATA_TG - anti-thyroglobulin antibodies; WBC - white blood cells; NEUT_ABS - absolute neutrophils count; MONO_ABS - absolute monocytes count; LYMPH_ABS - absolute lymphocytes count; MPV - mean platelet volume; PCT - plateletcrit; PDW - platelet distribution width; PLCR - platelet large cell ratio; PLT - platelet count; RDW - red cell distribution width.

Inspection of the network topology identified several densely connected regions corresponding to coherent physiological processes, whereas relatively few direct conditional dependencies were observed between biologically unrelated biomarkers. This organization suggests that the inferred network captures biologically meaningful conditional relationships rather than reflecting nonspecific correlation structure.

Overall, the reconstructed network demonstrated a modular yet integrated organization, supporting the hypothesis that PCOS is characterized by coordinated interactions among multiple physiological systems rather than by isolated abnormalities within individual endocrine or metabolic pathways.

### 3.3 Central biomarkers

Node-centrality analysis identified a subset of biomarkers occupying prominent positions within the inferred PCOS network. The highest overall node strength was observed for WBC, followed by total cholesterol, P-LCR, PCT, MPV, platelet count, TG, SHBG, HDL-C, and fasting insulin. These findings indicate that both hematological and metabolic biomarkers contributed substantially to the overall connectivity of the reconstructed network.

The strongest centrality signal was observed for WBC (strength = 1.456), suggesting that leukocyte-related variation represented one of the most highly connected components of the inferred biological architecture. Several platelet-related indices, including P-LCR, PCT, MPV, and platelet count, also ranked highly by node strength, indicating the presence of a densely connected hematological subnetwork.

Among metabolic biomarkers, TCHOL exhibited the second-highest node strength (strength = 1.196), whereas TG and HDL-C also occupied leading positions within the centrality ranking. Together, these findings indicate that lipid metabolism constitutes one of the principal organizational components of the inferred PCOS network.

Although SHBG and fasting insulin were not the highest-ranking nodes according to conventional centrality measures, both demonstrated consistently high overall connectivity while also participating in numerous inter-domain interactions. This distinction suggests that conventional node centrality and bridge centrality capture complementary aspects of network organization.

Shortest-path-based metrics further supported the importance of metabolic and hematological biomarkers. Triglycerides, PCT, GLU120, and absolute neutrophil count demonstrated high betweenness centrality, indicating that these biomarkers frequently participated in shortest communication paths connecting different regions of the network (Figure 3).

**Figure 3.**
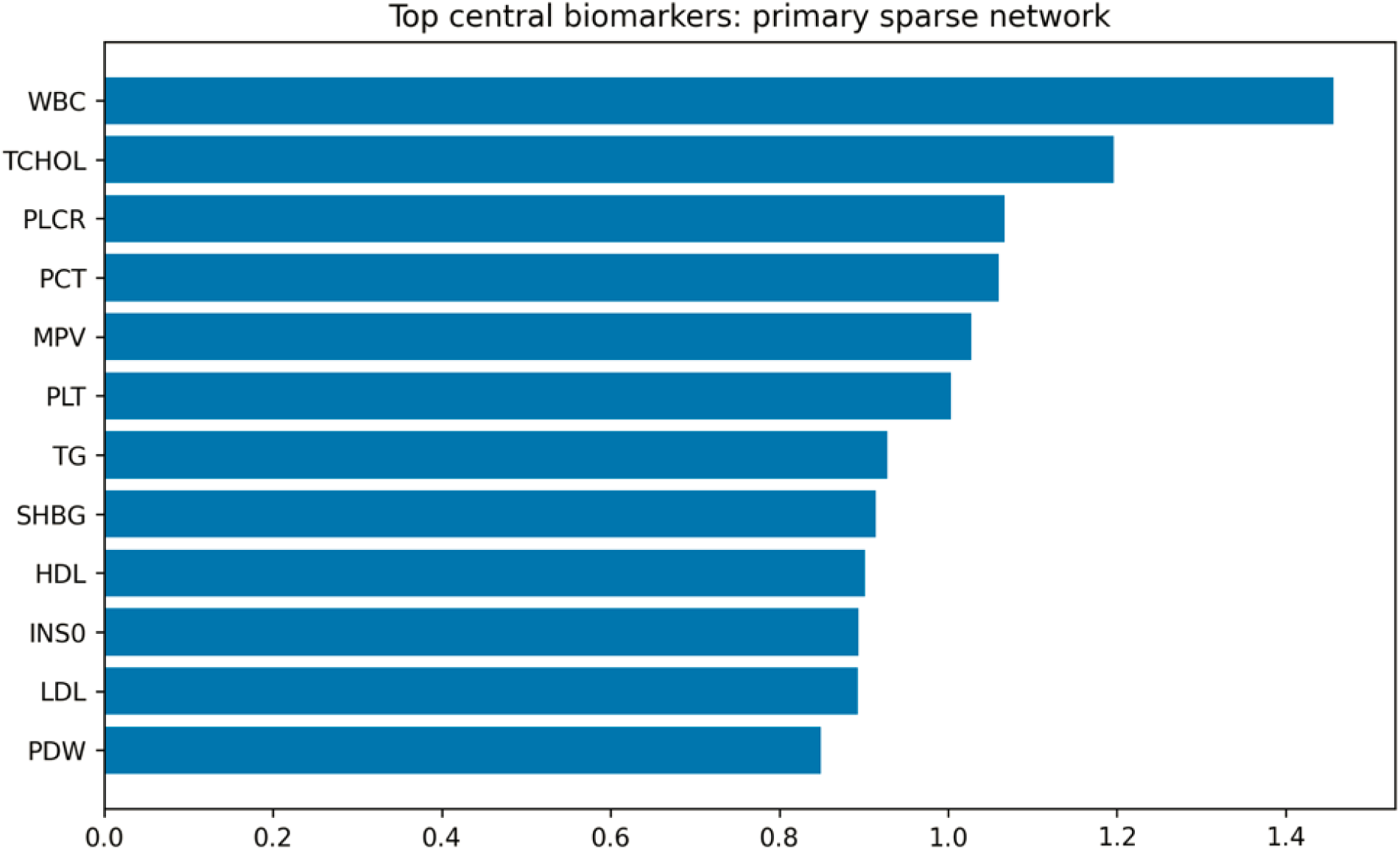
Node centrality analysis of the endocrine–metabolic biomarker network in women with polycystic ovary syndrome Legend: WBC - white blood cell count; TCHOL - total cholesterol; PLCR - platelet large cell ratio; PCT - plateletcrit; MPV - mean platelet volume; PLT - platelet count; TG - triglycerides; SHBG - sex hormone-binding globulin; HDL - high-density lipoprotein; INS0 - fasting insulin; LDL - low-density lipoprotein; PDW - platelet distribution width.

**Figure 4.**
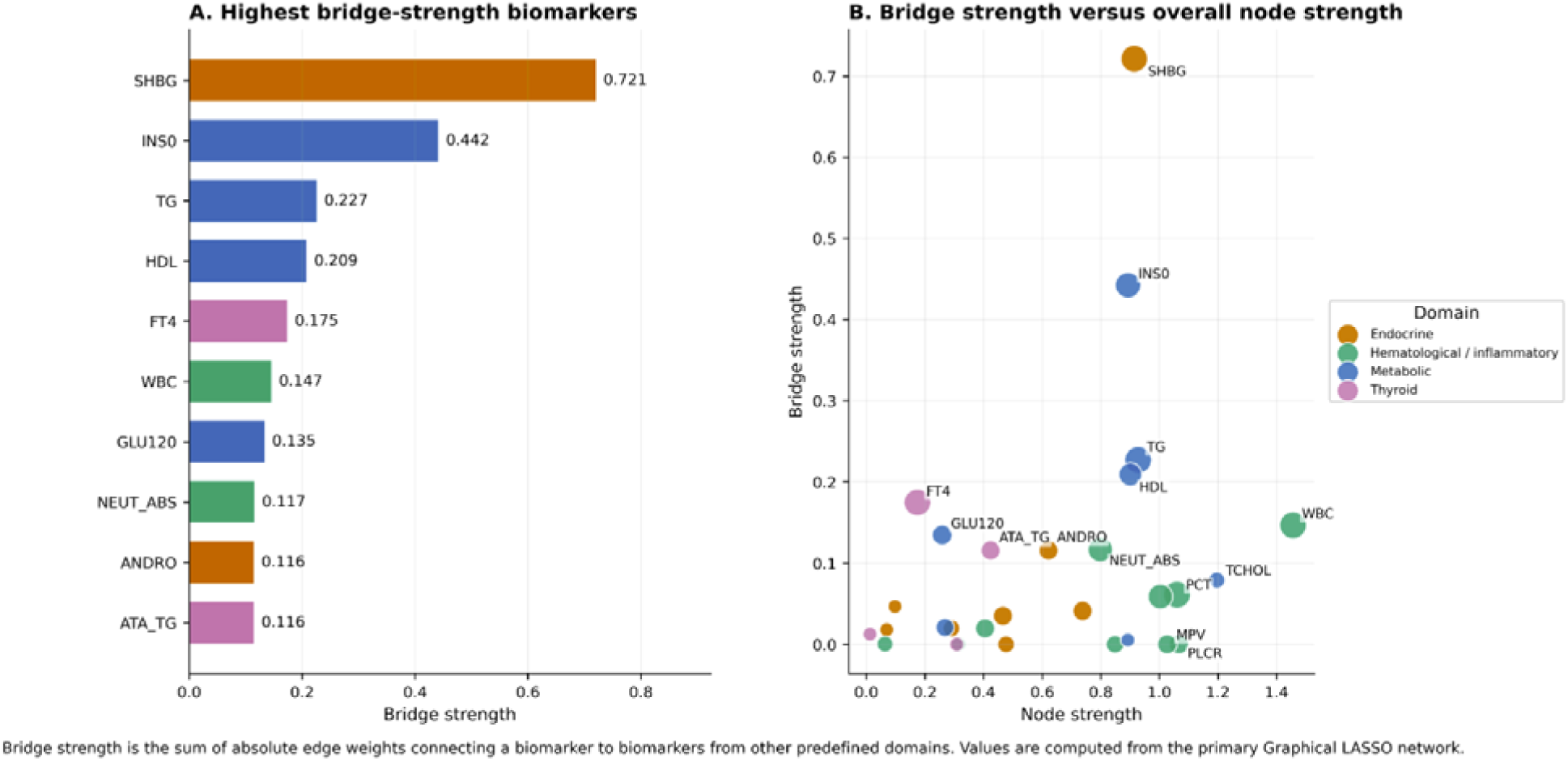
Bridge biomarkers integrating biological domains in the PCOS endocrine–metabolic network Legend: SHBG - sex hormone-binding globulin; INS0 - fasting insulin; TG - triglycerides; HDL - high density lipoprotein cholesterol; FT4 - free thyroxine; WBC - white blood cells; GLU120 - glucose after OGTT; NEUT_ABS - absolute neutrophils count; ANDRO - androstendione; ATA_TG - anti-thyroglobulin antibodies.

**Figure 5.**
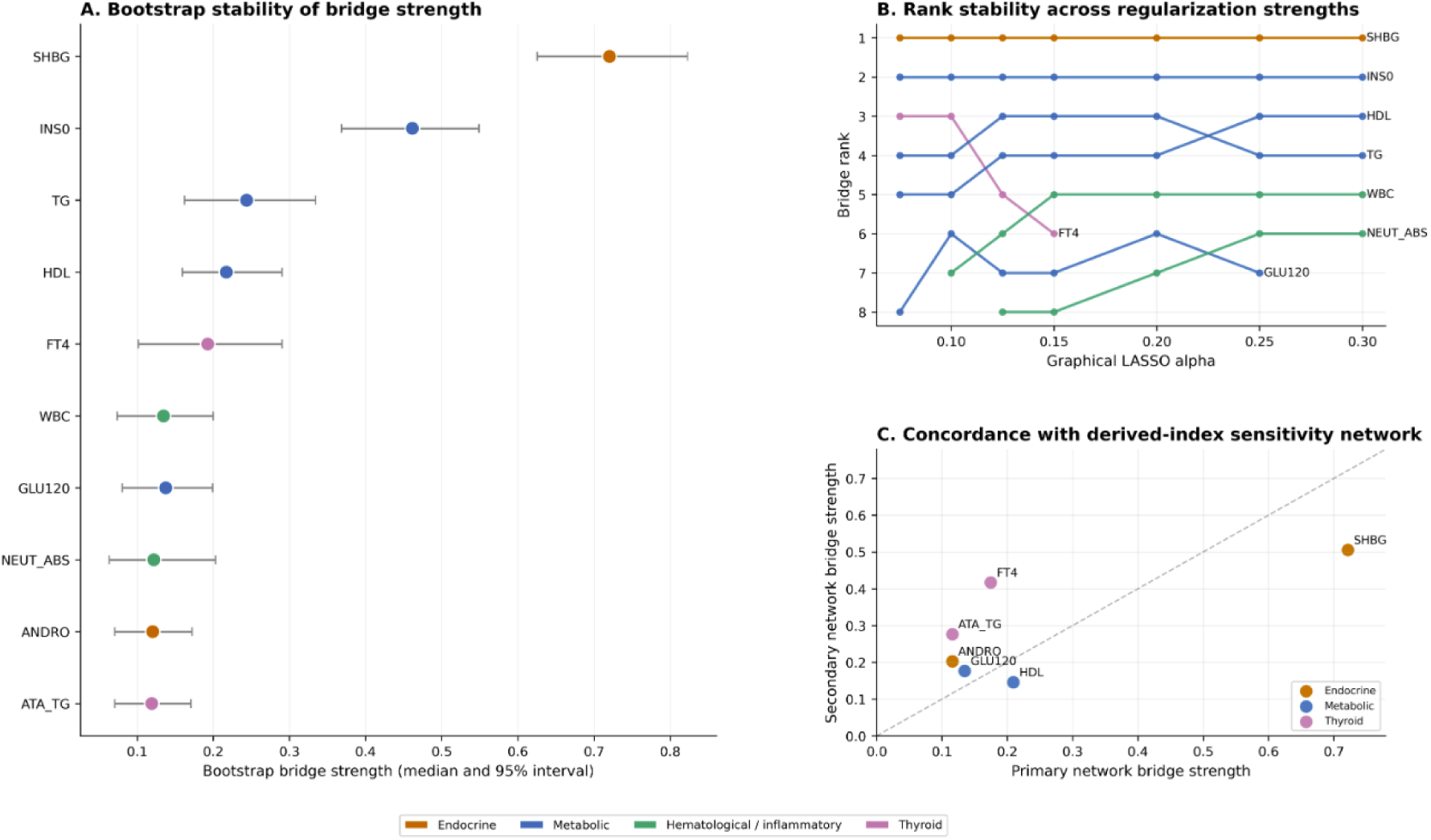
Robustness and stability of bridge biomarkers across complementary sensitivity analyses Legend: SHBG - sex hormone-binding globulin; INS0 - fasting insulin; TG - triglycerides; HDL - high density lipoprotein cholesterol; FT4 - free thyroxine; WBC - white blood cells; GLU120 - glucose after OGTT; NEUT_ABS - absolute neutrophils count; ANDRO - androstendione; ATA_TG - anti-thyroglobulin antibodies.

Collectively, these findings indicate that conventional node-centrality measures primarily identified biomarkers organizing local physiological modules, whereas the integration of distinct biological domains required dedicated bridge-centrality analysis. This observation motivated the subsequent evaluation of bridge biomarkers linking endocrine, metabolic, hematological and inflammatory, and thyroid systems.

### 3.4 Bridge biomarkers

Bridge-centrality analysis identified a limited subset of biomarkers preferentially connecting the predefined endocrine, metabolic, hematological and inflammatory, and thyroid domains, supporting the hypothesis that the biological organization of PCOS is coordinated by a relatively small number of inter-domain connectors rather than by uniformly distributed network connectivity.

Among all biomarkers included in the primary network, SHBG demonstrated the highest bridge strength, identifying it as the principal connector linking endocrine and metabolic regulation. Unlike conventional node-centrality measures, bridge strength specifically quantified the capacity of individual biomarkers to connect otherwise distinct physiological systems, highlighting the unique integrative role of SHBG within the reconstructed network.

The second highest bridge-centrality value was observed for fasting insulin (INS0), which linked glucose metabolism with endocrine and hematological processes. Additional highly ranked bridge biomarkers included triglycerides (TG) and HDL cholesterol, further emphasizing the central role of lipid metabolism in coordinating interactions between biological domains.

Several additional biomarkers—including FT4, WBC, GLU120, and absolute neutrophil count (NEUT_ABS)—also demonstrated relatively high bridge-centrality values. These biomarkers suggest potential interactions involving thyroid physiology, hematological regulation, and post-load glucose metabolism. However, because their biological roles as inter-domain connectors remain less well established, these findings should be regarded as exploratory and hypothesis-generating.

Similarly, androstenedione (ANDRO) and anti-thyroglobulin antibodies (ATA_TG) ranked among the higher bridge biomarkers, suggesting additional links between androgen metabolism, thyroid autoimmunity, and the broader endocrine–metabolic network. These observations warrant validation in independent cohorts before definitive biological conclusions can be drawn.

Importantly, several biomarkers exhibiting high bridge strength were not among the most central nodes according to conventional node-centrality measures. Conversely, biomarkers with high overall connectivity frequently remained confined to interactions within a single biological domain. These findings demonstrate that node centrality and bridge centrality describe complementary but biologically distinct properties of network organization.

Overall, bridge-centrality analysis indicates that the endocrine–metabolic architecture of PCOS is primarily coordinated by a small group of biologically plausible endocrine–metabolic connectors—particularly SHBG, fasting insulin, triglycerides, and HDL cholesterol—while additional bridge signals involving FT4, WBC, GLU120, NEUT_ABS, ANDRO, and ATA_TG provide exploratory evidence supporting broader systems-level integration.

### 3.5 Robustness analyses

The robustness of the inferred endocrine–metabolic network was evaluated through a series of predefined analyses addressing the principal sources of analytical uncertainty, including missing-data handling, network regularization, bootstrap resampling, and alternative network specifications.

Reconstruction of the network using the predefined complete-case dataset (n = 885 women with no missing values among the 29 primary biomarkers, 68.8% of the primary cohort) produced a global network topology that was largely concordant with the primary median-imputed network. The complete-case network retained 66 edges at a slightly higher regularization parameter (α = 0.15 vs. 0.125 in the primary network), with 90.4% of primary-network edges preserved (Jaccard similarity = 0.90) and high rank concordance for both node strength (Spearman ρ = 0.98) and bridge strength (Spearman ρ = 0.94). SHBG, fasting insulin, triglycerides, and HDL cholesterol occupied an identical top-four bridge-centrality ranking in both analyses; among the additional, lower-ranking bridge signals, FT4 and 2-hour plasma glucose in the primary network were replaced by androstenedione and anti-thyroglobulin antibodies in the complete-case network, biomarkers that had already been flagged as exploratory bridge signals in the primary analysis.

Similarly, variation of the Graphical LASSO regularization parameter across predefined neighboring values resulted in modest changes in network sparsity without materially altering the biological interpretation. The principal endocrine, metabolic, hematological and inflammatory, and thyroid domains remained interconnected through the same dominant bridge biomarkers.

Across these alternative regularization strategies, SHBG, fasting insulin (INS0), triglycerides (TG), HDL cholesterol, and FT4 consistently occupied the highest bridge-centrality positions. Additional biomarkers—including WBC, GLU120, NEUT_ABS, ANDRO, and ATA_TG—also remained reproducibly represented among the higher-ranking bridge biomarkers, although their biological interpretation was considered more exploratory.

Bootstrap resampling further demonstrated the stability of bridge-centrality estimates. The highest-ranking bridge biomarkers identified in the primary analysis remained consistently among the leading bridge nodes throughout bootstrap iterations, indicating that their identification was not driven by individual observations or sampling variability. Conventional node-centrality measures likewise demonstrated limited variability across bootstrap samples, supporting the robustness of the inferred network topology.

Complementary sensitivity analyses incorporating predefined derived biomarkers modestly increased local connectivity within metabolic and hematological domains, as expected from deterministic mathematical relationships. Nevertheless, these modifications did not materially alter the overall systems-level organization of the network or the identification of the principal bridge biomarkers.

Collectively, the robustness analyses demonstrate that the principal findings of this study were reproducible across multiple predefined analytical scenarios and were not dependent on a particular missing-data strategy, regularization parameter, or network specification. The consistent identification of SHBG, fasting insulin, triglycerides, HDL cholesterol, and FT4 as the leading bridge biomarkers supports the stability of the inferred endocrine–metabolic network architecture, whereas additional signals involving WBC, GLU120, NEUT_ABS, ANDRO, and ATA_TG provide complementary evidence of broader biological integration that merits further investigation.

### 3.6 Sensitivity analyses

To determine whether the principal biological conclusions depended on the predefined network specification, a series of complementary sensitivity analyses was performed using alternative biomarker sets and network configurations.

First, the primary network consisting exclusively of biologically independent laboratory biomarkers was compared with an expanded network incorporating clinically established derived indices, including composite endocrine, metabolic, and hematological variables. Inclusion of these derived biomarkers increased local network density, particularly within metabolic and hematological domains, reflecting expected mathematical relationships between primary measurements and their derived indices.

Despite these local structural changes, the overall organization of the network remained largely unchanged, and the same physiological domains remained interconnected. However, not all principal bridge biomarkers retained their dominant positions within the expanded network: SHBG, FT4, and HDL cholesterol remained among the highest-ranking bridge biomarkers, whereas fasting insulin (INS0) and triglycerides (TG) dropped substantially in bridge-centrality rank once mathematically derived indices were added. Because INS0 and TG are direct components of several of these derived indices (e.g., HOMA-IR, TyG index, TG/HDL-C ratio), this attenuation most likely reflects redundancy with their own derived counterparts within the expanded network rather than a loss of biological relevance.

Second, complementary analyses using alternative network specifications demonstrated that the principal biological interpretation was independent of the exact network composition. Although additional biomarkers introduced new local conditional dependencies, they did not alter the overall systems-level organization linking endocrine, metabolic, hematological and inflammatory, and thyroid domains.

Importantly, biomarkers identified as robust bridge biomarkers in the primary analysis - particularly SHBG and HDL cholesterol—remained consistently among the highest-ranking bridge nodes across predefined sensitivity analyses, including bootstrap resampling, the complete-case network, and alternative regularization parameters. FT4 also remained among the leading bridge biomarkers, suggesting a reproducible but more cautiously interpreted thyroid-related connector signal. INS0 and TG were robustly reproduced across bootstrap resampling, the complete-case network, and alternative alpha values, but ranked lower once mathematically derived indices were added to the network, consistent with redundancy with their own derived counterparts. Lower-ranking bridge biomarkers, including WBC, GLU120, NEUT_ABS, ANDRO, and ATA_TG, showed supportive but more exploratory evidence of cross-domain integration.

These findings indicate that the principal biological conclusions were not driven by the specific selection of biomarkers included in the primary network. Instead, the identification of a limited set of biomarkers coordinating communication between endocrine, metabolic, hematological and inflammatory, and thyroid physiology represented a reproducible characteristic of the inferred biological architecture.

Taken together with the robustness analyses, these results demonstrate that the systems-level organization of the PCOS biomarker network remained stable across alternative analytical assumptions, supporting the biological validity and reproducibility of the identified bridge biomarkers.

## 4. Discussion

### 4.1 Principal findings

The present study demonstrates that the biological organization of PCOS is best understood as an integrated network of interacting physiological systems rather than as a collection of isolated endocrine or metabolic abnormalities. Using a predefined Graphical LASSO framework, we reconstructed the conditional dependency architecture linking endocrine, metabolic, hematological and inflammatory, and thyroid biomarkers, revealing a sparse but highly coordinated biological network organized around a limited number of reproducible bridge biomarkers.

The principal finding of this study is that biomarkers contributing most strongly to the integration of biological domains were not necessarily those exhibiting the highest overall network connectivity. Conventional node-centrality analysis primarily identified biomarkers organizing local physiological modules, whereas bridge-centrality analysis revealed a distinct subset of biomarkers preferentially connecting endocrine, metabolic, hematological and inflammatory, and thyroid systems. This distinction highlights that overall network influence and inter-domain integration represent complementary but biologically different properties of the reconstructed PCOS network.

Among the identified bridge biomarkers, SHBG, fasting insulin, triglycerides, and HDL cholesterol emerged as the most biologically plausible endocrine–metabolic integrators. Additional bridge signals involving FT4, WBC, GLU120, NEUT_ABS, androstenedione, and anti-thyroglobulin antibodies suggested broader interactions between thyroid function, glucose metabolism, hematological regulation, and androgen biology. However, these latter findings should be interpreted as exploratory until confirmed in independent study populations.

Importantly, the inferred network architecture demonstrated a high degree of analytical stability. The principal bridge biomarkers remained consistently identified across complementary analyses evaluating missing-data handling, Graphical LASSO regularization, bootstrap resampling, and alternative network specifications. This reproducibility indicates that the observed systems-level organization was not driven by individual analytical assumptions but instead reflects robust structural characteristics of the underlying biological network.

Our findings extend the current understanding of PCOS by shifting the analytical focus from individual laboratory abnormalities toward the organization of interactions between physiological systems. Previous studies have predominantly investigated endocrine dysfunction, insulin resistance, dyslipidemia, inflammation, or thyroid abnormalities separately. Network-based approaches have previously been applied to PCOS at the transcriptomic level, for example through gene co-expression network analysis of granulosa cell expression data (18), but conditional-dependency networks constructed from routine clinical laboratory biomarkers remain comparatively unexplored. In contrast, the present network-based approach suggests that the biological complexity of PCOS arises from coordinated interactions among these systems rather than from isolated alterations within any single pathway.

Collectively, these findings support a systems biology model of PCOS in which disease heterogeneity emerges from the architecture of interactions linking multiple physiological domains. Within this framework, bridge biomarkers provide information that cannot be obtained from conventional analyses of individual biomarkers alone and may offer a more informative representation of the biological organization underlying PCOS.

### 4.2 Biological interpretation of bridge biomarkers

The identification of bridge biomarkers provides a systems-level perspective on PCOS by highlighting laboratory variables that preferentially connect distinct physiological domains rather than acting solely within individual biological pathways. Unlike conventional biomarkers, which are typically interpreted in isolation, bridge biomarkers reflect the integration of multiple regulatory systems and therefore offer insight into the biological architecture underlying disease complexity.

The highest bridge strength was observed for SHBG, identifying it as the principal integrator of the reconstructed endocrine–metabolic network. This finding is biologically plausible because SHBG occupies a central position at the interface between androgen availability, hepatic metabolism, and insulin action. Although traditionally regarded as a marker of hyperandrogenism, circulating SHBG concentrations are also strongly influenced by insulin resistance, adipose tissue dysfunction, hepatic metabolic activity, and chronic low-grade inflammation.(13) Consequently, SHBG is uniquely positioned to reflect coordinated regulation across multiple physiological systems rather than isolated endocrine function. Our network analysis extends this concept by demonstrating that SHBG preferentially connects distinct biological domains, supporting its role as a systems-level integrator within PCOS.

Fasting insulin (INS0) emerged as the second principal bridge biomarker, reinforcing the central role of insulin resistance in the pathophysiology of PCOS. Hyperinsulinemia influences ovarian steroidogenesis, suppresses hepatic SHBG synthesis, modifies lipid metabolism, and interacts with inflammatory signaling pathways.(13) Rather than representing only a metabolic abnormality, fasting insulin appears to function as a biological link between endocrine regulation and broader metabolic homeostasis. The prominent bridge position of fasting insulin within the reconstructed network is therefore consistent with its established role as one of the major drivers of PCOS pathophysiology.

The identification of triglycerides and HDL cholesterol among the highest-ranking bridge biomarkers further emphasizes the importance of lipid metabolism within the endocrine–metabolic network. Dyslipidemia in PCOS is increasingly recognized as a manifestation of systemic metabolic dysfunction rather than an isolated cardiovascular risk factor.(14) Their bridge-centrality positions suggest that lipid metabolism may participate in coordinating communication between endocrine and metabolic processes, extending beyond its traditional role as a downstream consequence of insulin resistance.

In addition to these principal endocrine–metabolic connectors, several biomarkers - including FT4, WBC, GLU120, and absolute neutrophil count - demonstrated reproducible but comparatively weaker bridge-centrality signals. These observations suggest that thyroid physiology, post-load glucose regulation, and hematological or inflammatory processes may also contribute to communication between biological domains. Elevated white blood cell and neutrophil counts have previously been reported as reproducible markers of chronic low-grade inflammation in PCOS (15,17). However, because these biomarkers have not previously been established as inter-domain connectors in PCOS, these findings should be interpreted cautiously and considered hypothesis-generating until validated in independent cohorts.

Similarly, the bridge-centrality positions of androstenedione and anti-thyroglobulin antibodies indicate potential interactions between androgen metabolism, thyroid autoimmunity, and the broader endocrine–metabolic network. Although these biomarkers did not emerge as the dominant integrators of network architecture, their reproducible identification across complementary analyses suggests that additional physiological pathways beyond classical insulin resistance and hyperandrogenism may contribute to the biological heterogeneity of PCOS. An increased prevalence of thyroid autoimmunity, including anti-thyroglobulin antibody positivity, has been reported in women with PCOS in systematic reviews and meta-analyses (16).

Taken together, the identified bridge biomarkers support the concept that PCOS is organized around a limited number of biological integrators linking endocrine regulation, glucose and lipid metabolism, hematological and inflammatory activity, and thyroid physiology. Rather than functioning as isolated indicators of individual pathological processes, these biomarkers appear to coordinate interactions across multiple physiological systems, providing a biologically coherent explanation for the multidimensional nature of PCOS.

### 4.3 Systems-level organization of PCOS

The present findings support the concept that PCOS should be viewed as a systems-level disorder emerging from coordinated interactions among multiple physiological domains rather than from isolated disturbances within individual endocrine or metabolic pathways. Although hyperandrogenism, insulin resistance, dyslipidemia, inflammation, and thyroid dysfunction have traditionally been investigated as separate components of PCOS, our network analysis indicates that these processes form an interconnected biological system characterized by structured conditional dependencies.

An important observation was that the reconstructed network exhibited a modular yet integrated architecture. Biomarkers clustered according to their predominant physiological functions, forming recognizable endocrine, metabolic, hematological and inflammatory, and thyroid domains. However, these domains were not biologically independent. Instead, they were linked through a limited number of bridge biomarkers that maintained communication between otherwise distinct physiological modules. Such an organization is characteristic of complex biological systems, in which overall function depends not only on the activity of individual components but also on the interactions connecting them.

This network perspective provides an alternative explanation for one of the defining characteristics of PCOS—its marked biological heterogeneity. Women with PCOS often present with similar clinical manifestations despite considerable differences in endocrine profiles, metabolic abnormalities, inflammatory status, and thyroid function. Rather than representing distinct and unrelated disease subtypes, these differences may reflect variation in the relative organization and strength of interactions among shared physiological systems. Consequently, the heterogeneity of PCOS may arise not only from differences in individual biomarker concentrations but also from differences in the architecture of biological interactions.

Our findings also suggest that conventional biomarker-based analyses may provide an incomplete representation of PCOS biology. Traditional statistical approaches primarily evaluate independent associations between individual variables and clinical outcomes, whereas network analysis characterizes the structure of relationships among biomarkers after accounting for the influence of all remaining variables. By focusing on conditional dependencies rather than isolated correlations, this approach enables identification of biological interactions that may remain undetected using conventional analytical strategies.

Importantly, the modular organization observed in the present study remained highly concordant across complementary robustness and sensitivity analyses, indicating that the inferred biological architecture was reproducible under multiple analytical assumptions. The preservation of both global network topology and the principal bridge biomarkers supports the interpretation that the reconstructed network captures stable features of PCOS biology rather than methodological artifacts introduced by model specification or missing-data handling.

Taken together, these observations support a systems biology framework in which PCOS is interpreted as an emergent property of interactions among endocrine, metabolic, hematological and inflammatory, and thyroid regulatory systems. Within this framework, the biological organization of the network may be more informative than isolated laboratory abnormalities, providing a conceptual basis for future studies investigating disease mechanisms, biological phenotyping, and systems-oriented approaches to PCOS.

### 4.4 Clinical implications

Although the present study was not designed to develop diagnostic or prognostic models, the reconstructed endocrine–metabolic network provides several clinically relevant insights into the biological organization of PCOS.

Current clinical evaluation of PCOS is largely based on the interpretation of individual laboratory parameters together with clinical manifestations of hyperandrogenism, ovulatory dysfunction, and ovarian morphology. While this approach remains fundamental for diagnosis, it does not fully capture the complex interactions between endocrine, metabolic, inflammatory, and thyroid systems that contribute to the biological heterogeneity of the syndrome. The present findings suggest that the clinical relevance of individual biomarkers may depend not only on their absolute concentrations but also on their position within the broader physiological network.

The identification of SHBG, fasting insulin, triglycerides, and HDL-C as the principal bridge biomarkers is particularly noteworthy because all four variables are routinely measured or readily obtainable during standard clinical assessment of women with PCOS. Rather than proposing these biomarkers as novel diagnostic tests, our results suggest that they may represent key indicators of communication between endocrine and metabolic systems. This network perspective may provide additional biological context for interpreting routinely available laboratory data without requiring the introduction of new or specialized assays.

The observed systems-level organization also has potential implications for biological phenotyping. Current PCOS phenotypes are primarily defined according to combinations of clinical and reproductive characteristics, whereas the reconstructed network suggests that women with similar clinical presentations may nevertheless differ in the organization of interactions among endocrine, metabolic, hematological and inflammatory, and thyroid domains. Future studies integrating network topology with detailed clinical phenotypes may therefore contribute to more biologically informed stratification of PCOS.

From a translational perspective, network analysis may also facilitate the identification of biomarkers that occupy strategically important positions within the biological system. Biomarkers functioning as inter-domain connectors may represent informative candidates for monitoring systemic responses to lifestyle interventions, pharmacological treatment, or longitudinal disease progression. However, these potential applications remain speculative and require prospective validation before any clinical implementation can be recommended.

Importantly, the present findings should not be interpreted as evidence that bridge biomarkers are superior to established diagnostic criteria or existing biochemical markers. Rather, they demonstrate that network analysis provides complementary information describing how routinely measured biomarkers interact within the broader biological system. Such systems-level information may ultimately support precision medicine approaches by improving the biological characterization of women with PCOS, but confirmation in independent cohorts and longitudinal studies will be essential before translation into clinical practice.

Overall, the present study illustrates how network-based analytical approaches can complement conventional endocrinological investigations by shifting the focus from isolated biomarkers toward the organization of biological interactions. This perspective may contribute to a more comprehensive understanding of PCOS and provide a conceptual framework for future studies investigating individualized disease mechanisms and systems-oriented patient stratification.

### 4.5 Strengths and limitations

The present study has several important strengths. First, to our knowledge, this is among the first studies to comprehensively characterize the biological architecture of PCOS using a network-based framework integrating endocrine, metabolic, hematological and inflammatory, and thyroid biomarkers within a single analytical model. Rather than focusing on isolated associations between individual laboratory variables, the study reconstructed the conditional dependency structure underlying routinely measured clinical biomarkers, providing a systems-level representation of PCOS biology.

Second, the analytical workflow was fully predefined and implemented within a reproducible computational pipeline. Variable selection, preprocessing procedures, missing-data handling, Graphical LASSO model selection, bridge-centrality analysis, and all robustness and sensitivity analyses were specified before biological interpretation. This strategy minimized investigator-dependent analytical decisions and reduced the risk of post hoc optimization or selective reporting.

Third, the primary findings were supported by multiple complementary robustness analyses, including comparison with complete-case reconstruction, alternative Graphical LASSO regularization parameters, bootstrap resampling, and alternative network specifications. The consistent identification of the principal bridge biomarkers across these independent analyses increases confidence that the observed network architecture reflects reproducible biological structure rather than analytical artifacts.

The study should also be interpreted in the context of several limitations. The cross-sectional design precludes inference regarding causal relationships between biomarkers or the temporal direction of interactions within the reconstructed network. Conditional dependencies identified by Gaussian graphical models describe statistical relationships after adjustment for the remaining variables but should not be interpreted as evidence of direct biological causation.

Second, although the study included a large, well-characterized cohort of women with PCOS, all participants originated from a single tertiary referral center. Consequently, the reconstructed network architecture may partly reflect characteristics of this specific clinical population, and external validation in independent cohorts representing different ethnicities, healthcare settings, and age groups will be necessary to establish generalizability.

Third, missing laboratory measurements are an inherent feature of retrospective clinical datasets. Although the primary analyses were performed using a predefined median-imputation strategy and supported by a complete-case sensitivity analysis retaining 68.8% of the cohort (n = 885), any imputation approach necessarily relies on assumptions regarding the underlying data structure. The substantial concordance between primary and complete-case analyses, particularly for the four leading bridge biomarkers (SHBG, INS0, TG, HDL-C), reduces this concern but does not eliminate it entirely, especially for lower-ranking, more exploratory bridge signals that were more sensitive to the missing-data handling strategy.

Finally, the biological domains used for bridge-centrality analysis were defined a priori according to current physiological knowledge. Although this approach enhances interpretability and prevents data-driven domain assignment, alternative biological classifications could produce modest differences in bridge-centrality rankings. Likewise, the identification of bridge biomarkers reflects the reconstructed network architecture within this specific analytical framework and should therefore be interpreted as evidence of biological organization rather than definitive proof of mechanistic interactions.

Despite these limitations, the combination of a large clinical cohort, predefined analytical strategy, rigorous robustness assessment, and comprehensive systems-level network analysis provides a solid methodological foundation for the present findings. We believe that the reconstructed endocrine–metabolic network offers a reproducible framework for investigating the biological organization of PCOS and establishes a basis for future longitudinal, multi-center, and mechanistic studies designed to validate and extend these observations.

## 5. Conclusions

This study demonstrates that the biological organization of polycystic ovary syndrome extends beyond isolated endocrine or metabolic abnormalities and is more appropriately understood as an integrated network of interacting physiological systems. Using a predefined Graphical LASSO framework, we reconstructed the conditional dependency architecture linking endocrine, metabolic, hematological and inflammatory, and thyroid biomarkers, revealing a sparse yet highly coordinated network organized around a limited number of reproducible bridge biomarkers.

Among the identified bridge biomarkers, SHBG, fasting insulin, triglycerides, and HDL-C emerged as the principal integrators of the endocrine–metabolic network, whereas additional bridge signals involving FT4, WBC, GLU120, NEUT_ABS, androstenedione, and anti-thyroglobulin antibodies suggest broader interactions among multiple physiological systems that warrant further investigation. Importantly, the reconstructed network architecture remained highly concordant across predefined robustness and sensitivity analyses, supporting the reproducibility of the principal findings.

Collectively, these findings support a systems biology perspective of PCOS in which disease heterogeneity arises from coordinated interactions among multiple biological domains rather than from isolated alterations in individual biomarkers. Network-based characterization of routinely measured laboratory variables provides complementary information beyond conventional biomarker analyses and offers a reproducible framework for future studies investigating biological phenotypes, disease mechanisms, and precision medicine approaches in PCOS.

## Data Availability

https://github.com/npiorkowska-science/pcos-network-architecture

## Declarations

### Data Availability

The clinical dataset analyzed during the current study is not publicly available because it contains sensitive patient information and institutional ethical restrictions prohibit unrestricted public release. De-identified data may be made available from the corresponding author upon reasonable request and subject to approval by the appropriate institutional and ethical authorities.

### Code Availability

The complete analytical workflow developed for this study, including Python scripts for data harmonization, preprocessing, missing-data handling, dimensionality reduction, clustering, stability assessment, cross-space agreement analysis, biological validation, sensitivity analyses, and figure generation, is openly available in the following GitHub repository:

https://github.com/npiorkowska-science/pcos-network-architecture

The repository also contains documentation describing the computational workflow, software dependencies, and instructions required to reproduce all analyses presented in this manuscript.

### Ethics Approval

This study was conducted in accordance with the Declaration of Helsinki and was approved by the Bioethics Committee of Wroclaw Medical University (approval no. 254/2021).

### Consent to Participate

Written informed consent was obtained from all participants prior to inclusion in the study.

### Consent for Publication

All authors have reviewed and approved the final version of the manuscript and consent to its publication.

### Author Contributions

Natalia Piórkowska: Conceptualization, methodology, software, formal analysis, investigation, data curation, visualization, writing – original draft, writing – review and editing.

Grzegorz Franik: Data curation.

Anna Bizoń: Writing – review and editing, supervision.

All authors read and approved the final manuscript.

## Acknowledgements

The authors thank all study participants and the clinical staff involved in patient recruitment, data collection, and laboratory diagnostics.

## Artificial Intelligence Disclosure

Artificial intelligence (AI)-assisted tools were used exclusively for the preparation of the graphical illustration presented in Figure 1. This figure was created solely to improve the visual presentation of the study workflow and does not contain AI-generated scientific content.

